# NLRP3 Inflammasome, NEK7 and Major Depressive Disorder

**DOI:** 10.1101/2022.07.16.22277717

**Authors:** Fatih Ozel, Bilge Targitay Ozturk, Tutku Yaras, Burcu Ekinci, Yavuz Oktay, Tunc Alkin, Elif Onur Aysevener, Nese Direk

## Abstract

**Background:** Association between inflammation and depression has been known for a long time. Activation of pro-inflammatory molecular complexes such as inflammasomes in depression was suggested as the most relevant hypothesis among many others. Psychological stress is considered to cause sterile inflammation through inflammasomes, and the NLRP3 inflammasome was proposed as a crucial molecule for the pro-inflammatory response in depression.

**Objective:** In the current study, we aimed to explore the relationship of NLRP3 inflammasome and its regulatory protein NEK7 with major depressive disorder in a drug naïve study sample.

**Methods:** In total 58 patients with major depressive disorder and 58 age and gender-matched healthy persons were included. The mRNA expressions of NLRP3, ASC, caspase-1 and NEK7 coding proteins were evaluated with quantitative PCR, plasma IL-1β levels were detected by ELISA.

**Results:** Patients with major depressive disorder had higher gene expressions of NLRP3 (p= 0.03) and ASC (p= 0.002) compared to healthy persons. Higher gene expressions of NLRP3 (OR= 1.17, 95% CI= 1.01, 1.37, p= 0.04), ASC (OR= 1.45, 95% CI= 1.15, 1.82, p= 0.002) and NEK7 (OR= 1.33, 95% CI= 1.08, 1.63, p= 0.007) were related to the increased likelihood of having major depressive disorder.

**Conclusion:** The results of this study support the role of NLRP3 inflammasome in the increased risk for major depressive disorder.

## 1. Introduction

Major depressive disorder (MDD) is a severe psychiatric disorder that is one of the leading causes of mortality and morbidity worldwide (Malhi and Mann, 2018). Despite mounting evidence, the etiology of MDD remains highly heterogeneous. Inflammatory processes seem to play an important role in the etiology of MDD, among many other contributing factors (Malhi and Mann, 2018). Increased levels of pro-inflammatory cytokines including IL-1, IL-6, and TNF-α in MDD have been identified before (Haapakoski et al., 2015; Howren et al., 2009; Liu et al., 2012), though the evidence is limited. Therefore, exploring specific inflammatory molecules in MDD is essential to shed light on the inconclusive findings.

Inflammasomes are complex molecules that control many aspects of the inflammatory response (Martinon et al., 2002). The NLRP3 inflammasome is the most studied inflammasome complex in humans and it has been linked to a variety of stress-related diseases. It consists of the sensory protein NLRP3, the adaptor protein ASC and the precursor protein pro-caspase-1 (Schroder et al., 2010). The NLRP3 inflammasome is activated by different stimuli such as ions, mitochondrial substances, reactive oxygen species, and fatty acids (Swanson et al., 2019). Psychological stress is one of the most prominent stimuli of the NLRP3 inflammasome (Iwata et al., 2013). The activation of the NLRP3 inflammasome complex results in the synthesis of IL-1β and IL-18 (Schroder et al., 2010).

The relationship between the NLRP3 inflammasome and MDD has primarily been investigated mostly in animal studies. Researchers showed increased mRNA and protein expressions of the NLRP3 inflammasome complex in animal models for depression (Kaufmann et al., 2017; Nishiguchi et al., 2021; Ślusarczyk et al., 2016; Xie et al., 2021; Zhang et al., 2014). Similarly, inhibition of the NLRP3 inflammasome was related to improved outcomes in depressive symptoms in animal studies (Guo et al., 2020; Zhang et al., 2015). Furthermore, fluoxetine, a well-known selective serotonin reuptake inhibitor, was shown to inhibit NLRP3 inflammasome formation (Du et al., 2016; Pan et al., 2014). In clinical studies of the association between NLRP3 and MDD, a few studies found that the subunits of the NLRP3 inflammasome complex were partially increased in individuals with MDD compared to healthy controls (Alcocer-Gómez et al., 2017, 2014; Taene et al., 2020).

Many identified biomolecules regulate the activation of the NLRP3 inflammasome complex (Kelley et al., 2019). NIMA-related kinase 7 (NEK7) is a central regulatory molecule of the NLRP3 inflammasome complex (Schroder et al., 2010; Shi et al., 2016). Furthermore, the fact that NEK7 is unrelated to the activation of other inflammasome complexes such as NLRC4 and AIM makes it more specific for NLRP3 (Kelley et al., 2019). However, the potential role of NEK7 in the pathogenesis of MDD has never been investigated in clinical settings so far.

As a result, the evidence for a link between MDD and inflammation is still unclear, with results from cytokine studies providing the majority of support. Even though the NLRP3 inflammasome may be a key molecule in the inflammatory response, the relationship between the NLRP3 inflammasome and MDD has only been studied in a few small clinical studies. The aim of this study was to demonstrate a connection between the NLRP3 inflammasome and MDD.

## 2. Methods

### 2.1. Study sample and design

This study was conducted in Dokuz Eylul University Hospital, Outpatient Psychiatry Unit between January 2018 and January 2019. The patient group included 58 individuals with MDD. All psychiatric diagnoses were made upon clinical interview which was performed by two experienced psychiatrists (FÖ, BTO) with the Structured Clinical Interview for the DSM-IV Axis I Disorders (SCID-I). Additionally, patients with MDD were assessed with Hamilton Depression Rating Scale (HDRS), the ones with a score of 18 or above were included in the patient group. The healthy control group consisted of 58 participants who were evaluated with SCID-I by the same psychiatrists (FÖ, BTO). After clinical interviews, the participants without diagnoses of MDD, dysthymia, bipolar and related disorders, schizophrenia and other disorders with psychosis, generalized anxiety disorder, panic disorder, post-traumatic stress disorder were included in the healthy control group. Both groups comprise the participants aged between 30 and 75.

The exclusion criteria for both groups included a history of cardiovascular, cerebrovascular or autoimmune disease, active cancer, pregnancy and breastfeeding during the assessment, use of immunomodulators, systemic and topical steroids, anti-aggregates and anti-coagulants, psychotropic drugs excluding benzodiazepines and anti-histamines in the previous month, mental retardation, presence of visual and hearing impairment, illiteracy, serious cognitive impairment and having current somatic symptoms related to infection.

The current study was conducted in accordance with the latest version of the Declaration of Helsinki and approved by the Clinical Research Ethical Committee of Dokuz Eylul University in December 2017 (Protocol number: 416-SBKAEK). After the nature of the procedures had been fully explained, written informed consent was obtained from all participants.

### 2.2. Measures of inflammatory parameters

Blood samples were collected from patients and healthy controls. To isolate plasma, 3-4 ml blood was centrifuged at 2200 rpm for 10 minutes in 15 ml centrifuge tubes. The top layer was made up of plasma, which was aliquoted as 500 ul in cryotubes and stored at -80 °C. ELISA tests were applied for Human IL-1β (BioLegend, #437004) according to the protocol.

Blood samples in EDTA tubes were used for peripheral blood mononuclear cell (PBMC) isolation. Six ml of fresh blood was used for every patient and control. Blood samples were taken into a 15 ml centrifuge tube and 1:1 ratio phosphate-buffered saline (PBS) was added and mixed. As ratio 1:1:1, the lymphocyte separation medium (Lonza, #BE17-829E) was added, and PBS and blood mixture were added onto this solution without mixing. Samples were centrifuged at 2200 rpm for 25 minutes (acceleration, break 0). After centrifugation PBMC layer was collected into a new 15 ml centrifuge tube and 2 times washed with PBS (1500 rpm, 10 min, accel, break 9). After that, PBMCs were dissolved in TRI reagent (Sigma, #T9424) and kept at -80 ° C. For RNA isolation, Direct-zol RNA Miniprep Plus Kit (Zymo, #R2072) was used and the kit protocol was followed. RNA samples were kept at - 80 ° C. After that, the First Strand cDNA Synthesis Kit (ProtoScript, #E6300) was used with these RNA samples and random primers. These prepared cDNAs were kept at -20 °C. Prepared cDNA samples were used for quantitative RT-PCR preparation. Samples were prepared by using GoTaq qPCR Master Mix (Promega, #A6001), same conditions were applied for all samples which were 8 ul of GoTaq qPCR Master Mix, 3 ul of cDNA, 0,5 ul of forward primer, 0,5 ul of reverse primer and 4 ul of MilliQ. Different primers were used; NLRP3 primers were 5’-GCAGCAAACTGGAAAGGAAG (forward) and 5’-CTTCTCTGATGAGGCCCAAG -3’ (reverse), ASC primers were 5’-AGTTTCACACCAGCCTGGAA - 3’ (forward) and 5’-TTTTCAAGCTGGCTTTTCGT -3’ (reverse), caspase-1 primers were 5’-CTCAGGCTCAGAAGGGAATG -3’ (forward) and 5’-CGCTGTACCCCAGATTTTGT -3’ (reverse), NEK7 primers were 5’-TTTACACTCCTGACAGCG -3’ (forward) and 5’-GCAACAGGAACTTTAGAACT -3’ (reverse), GAPDH primers were 5’-TGCACCACCAACTGCTTAGC -3’ (forward) and 5’-GGCATGGACTGTGGTCATGAG -3’ (reverse). All measurements were done by using Roche 480 Light Cycler. All samples were prepared in triple replicates. For every gene, CT value was determined according to the housekeeping gene (GAPDH) and every reaction was normalized (Delta CT values were determined). Relative gene expression was calculated for each gene and sample.

### 2.3. MDD assessment

The clinical diagnoses were made with SCID-I as mentioned above. Detailed explanations for the user guide of SCID-I could be found elsewhere (First, 1997) and SCID-I was validated in the Turkish population before (Corapçıoğlu A., 1999). The severity of depression was assessed with the structured interview guide of the seasonal affective version of HDRS. This scale consists of 29 items with eight items are targeted to determine atypical features of MDD. To calculate the atypia index, the total score of eight items for the atypical features is divided by the total score of the whole scale (Williams et al., 1992). The HDRS is a Likert-type scale in which higher scores reflect more severe depression. The structured interview guide of the seasonal affective version of HDRS was also validated in Turkey (Aydemir et al., 2006).

### 2.4. Statistical analysis

In the descriptive analysis, means and standard deviations were presented for the numeric variables and proportions were presented for categorical variables. To assess the differences among patient and healthy control groups, a Student’s t-test was used for continuous variables, a chi-square test was used for categorical variables.

The associations between gene expressions of NLRP3 inflammasome complex, NEK7, level of IL-1β and MDD were assessed with the logistic regression analyses. Analyses were performed in two different models. In model 1, we adjusted analyses for age and gender. In model 2, we further adjusted analyses for education, household income per capita and body mass index. In the MDD group, correlations between gene expression of NLRP3, ASC, caspase-1, NEK, level of IL-1β and HDRS score, atypia index were tested with a Pearson correlation test.

All analyses were performed with IBM SPSS version 24. The statistical significance threshold was 0.05 for all analyses.

## 3. Results

Table 1 shows the descriptive characteristics of the groups. The mean age was 43.62 years (SD=10.27) for the MDD group and 44.14 years (SD=10.29) for the control group (p>0.05). Both groups were comparable in terms of gender. Household income per capita in the MDD group (mean=1597.6 Turkish Lira, SD=1031) was significantly lower than in the control group (mean=2377.5 Turkish Lira, SD=1440.5). Among MDD patients, 29 had melancholic and 6 had atypical subtype of depression. Half of the MDD group had their first episode of depression in their lifetime. The expression level of the NLRP3 was higher in the MDD group (mean=32.3, SD=2.73) than in the healthy control group (mean=31.21, SD=2.8). Similarly, ASC was expressed more in the MDD group (mean=33.83, SD=1.84) than in healthy controls (mean=32.65, SD=2.22). There were no significant differences in terms of caspase-1 and NEK7 expressions and IL-1β levels between the two groups.

**Table 1:** Descriptive characteristics of MDD and healthy control groups

| | MDD<br>(n=58) | Controls<br>(n=58) | t (df) / $\chi^2$ (df) | p value |
| --- | --- | --- | --- | --- |
| Age, years, mean (SD) | 43.62 (10.27) | 44.14 (10.29) | 0.27 (114) | 0.79 |
| Women, N (%) | 46 (%79.3) | 46 (%79.3) | 0 (1) | 1.00 |
| Education, years, mean (SD) | 11.10<br>(4.57) | 11.31<br>(5.18) | 0.23 (114) | 0.82 |
| Household income per capita,<br>TRY, mean (SD) | 1597.64<br>(1031.01) | 2377.50<br>(1440.45) | 3.41<br>(104.76) | 0.001 |
| Smoking status, N (%) |  |  | 1.15 (2) | 0.56 |
| Never | 20 (%34.5) | 21 (%36.2) |  |  |
| Former | 11 (%19.0) | 15 (%25.9) |  |  |
| Current | 27 (%46.6) | 22 (%37.9) |  |  |
| BMI, kg/m<sup>2</sup>, mean, (SD) | 27.24 (6.05) | 26.20 (5.54) | -0.96 (114) | 0.34 |
| Hypertension, N (%) | 12 (%20.7) | 13 (%22.4) | 0.05 (1) | 0.82 |
| Diabetes, N (%) | 5 (%8.6) | 6 (%10.3) | 0.10 (1) | 0.75 |
| NLRP3, mean (SD) | 32.30 (2.73) | 31.21 (2.80) | -2.11 (114) | 0.03 |
| ASC, mean (SD) | 33.83 (1.84) | 32.65 (2.22) | -3.12 (114) | 0.002 |
| Caspase-1, mean (SD) | 31.63 (2.11) | 31.35 (2.25) | -0.69 (114) | 0.49 |
| NEK7, mean (SD) | 33.15 (2.20) | 32.42 (2.44) | -1.68 (114) | 0.09 |
| IL-1β, pg/ml, mean (SD) | 29.24 (15.40) | 30.98 (15.32) | 0.61 (114) | 0.54 |

SD: standart deviation, df: degrees of freedom, TRY: Turkish lira, BMI: body mass index

Logistic regression analyses showed that higher gene expression of NLRP3 (OR=1.17, 95% CI=1.01, 1.37, p=0.04), ASC (OR=1.45, 95% CI=1.15, 1.82, p=0.002) and NEK7 (OR=1.33, 95% CI=1.08, 1.63, p=0.007) were associated with increased likelihood of having MDD in fully adjusted models (Table 2). No significant relations of expressions of caspase-1 and IL-1β level with MDD. In MDD group, no significant correlations were found between inflammasome complex, IL-1β levels and HDRS scores and atypia index (Table 3).

**Table 2:**
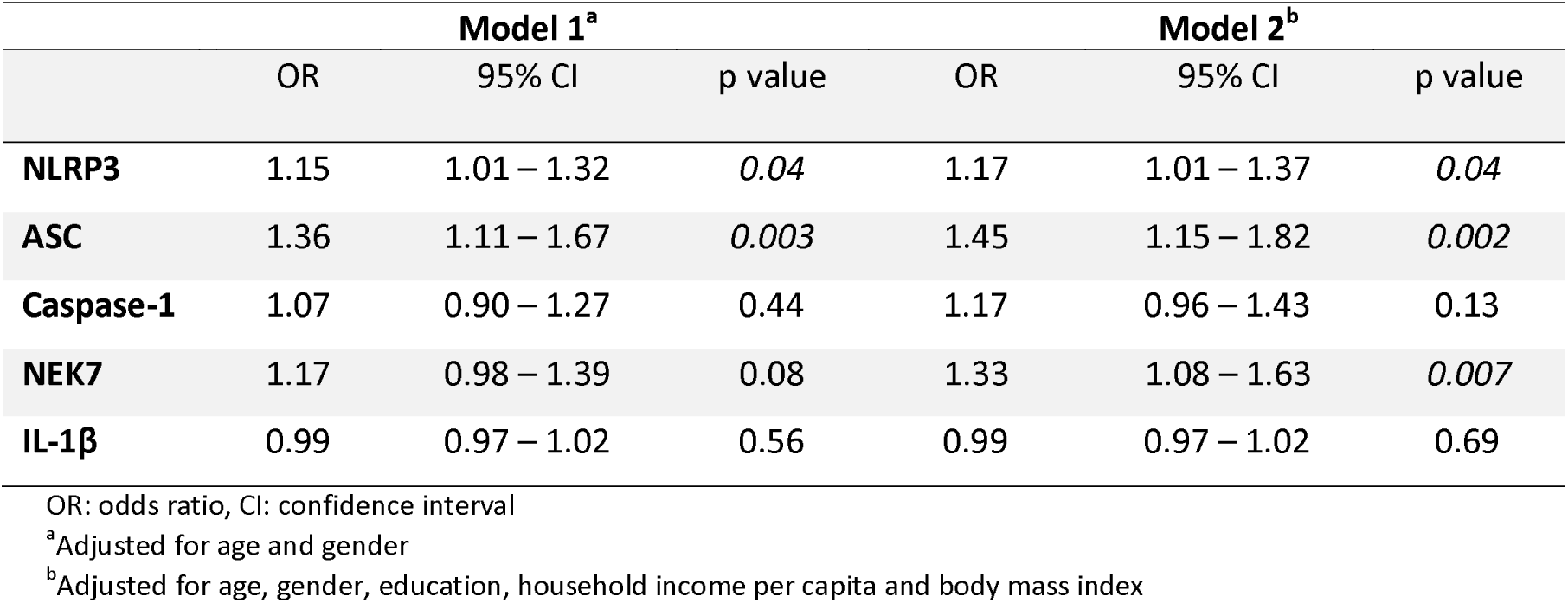
Associations between inflammatory parameters and MDD

|  | Model 1<sup>a</sup> |  |  | Model 2<sup>b</sup> |  |  |
| --- | --- | --- | --- | --- | --- | --- |
|  | OR | 95% CI | p value | OR | 95% CI | p value |
| NLRP3 | 1.15 | 1.01 – 1.32 | 0.04 | 1.17 | 1.01 – 1.37 | 0.04 |
| ASC | 1.36 | 1.11 – 1.67 | 0.003 | 1.45 | 1.15 – 1.82 | 0.002 |
| Caspase-1 | 1.07 | 0.90 – 1.27 | 0.44 | 1.17 | 0.96 – 1.43 | 0.13 |
| NEK7 | 1.17 | 0.98 – 1.39 | 0.08 | 1.33 | 1.08 – 1.63 | 0.007 |
| IL-1β | 0.99 | 0.97 – 1.02 | 0.56 | 0.99 | 0.97 – 1.02 | 0.69 |

OR: odds ratio, CI: confidence interval
<sup>a</sup>Adjusted for age and gender
<sup>b</sup>Adjusted for age, gender, education, household income per capita and body mass index

**Table 3:** Correlations between inflammatory parameters and depressive measures

|  | HDRS score | Atypia Index |
| --- | --- | --- |
| **NLRP3** | 0.21 | -0.14 |
| **ASC** | 0.02 | 0.05 |
| **Caspase-1** | -0.12 | 0.03 |
| **NEK7** | -0.07 | 0.14 |
| **IL-1β** | 0.13 | 0.01 |

Pearson correlation coefficients (r) are presented.
HDRS: Hamilton Depression Rating Scale
\*p<0.05

## 4. Discussion

In this clinical case-control study, mRNA expressions of NLRP3, ASC and NEK7 were associated with MDD. The components of NLRP3 and IL-1β were not associated with depression severity and atypia index.

The association between MDD and inflammation was first proposed 30 years ago (Smith, 1991); however, the studies that scrutinize NLRP3 inflammasome in this context accelerated in the last decade. The common consensus from these studies is that a range of molecules and psychological stress can stimulate the NLRP3 inflammasome and this may play a role in the pathogenesis of MDD (Franklin et al., 2018). In previous clinical studies, higher gene expressions of NLRP3 and caspase-1 in depressive patients than in healthy individuals were found (Alcocer-Gómez et al., 2017, 2014; Taene et al., 2020) which are overall in line with our findings. However, we did not find any associations between caspase-1 and MDD in our study. Although caspase-1 is produced upon the activation of the NLRP3 inflammasome, there are many other inflammasome complexes (i.e. NLRP1, NLRC4, AIM2, pyrin) that are responsible for the maturation of caspase-1 (Mathur et al., 2017). The fact that no study, including our study, explored other related pathways for caspase-1 might be an explanation for the conflicting findings. Caspase-1 should not be acknowledged as a direct indicator of NLRP3 activation and results need to be interpreted carefully. Regarding ASC, there is no information on the link between the gene expression of ASC and MDD in the previous clinical studies. However, outcomes of animal studies showed the connection between stress exposure and increased expression of ASC (Iwata et al., 2016; Ślusarczyk et al., 2016). In a similar manner, our finding on the positive relationship between gene expression of ASC and MDD supports the idea of NLRP3 activation in MDD.

NLRP3 inflammasome is activated by many factors including psychological stress (Iwata et al., 2013), which is also a significant risk factor MDD. Physical or psychological stress can cause the release of endogenous signal molecules named damage-associated molecular patterns (DAMPs) (Chen and Nuñez, 2010). These molecules interact with cellular receptors and this might result in the activation of the NLRP3 inflammasome (Chen and Nuñez, 2010). Pro-inflammatory cytokines which are produced upon the activation of inflammasomes may play role in the pathogenesis of MDD.

NEK7 is responsible for the regulation of centrosomes and microtubules, mitochondrial functions, repair of DNA damage (Liu et al., 2020). Upon the dysregulated homeostasis, NEK7 is produced in cells and triggers the pro-inflammatory response including the activation of NLRP3 (He et al., 2016; Saloura et al., 2015). To our best knowledge, there is no study that investigated the role of NEK7 in MDD in humans. The findings from this study on the relationship between NEK7 and MDD underline the importance of NLRP3 in MDD pathogenesis since NEK7 is known to activate only NLRP3 among many other inflammasome complexes (Kelley et al., 2019). Still, contributions of other potential biological mechanisms to MDD through NEK7 are required to be explored widely.

Even though the pro-inflammatory state in MDD is accepted to a large extent, the evidence for IL-1β is still inconclusive. Many recent meta-analyses could not find any difference between depressive patients and healthy controls regarding IL-1β (Goldsmith et al., 2016; Köhler et al., 2017). Similarly, we did not show any relation between MDD and the levels of IL-1β in this study. There may be some explanations for these heterogeneous findings. First, the pro-inflammatory response is thought to become more prominent with increasing age and late-life depression is seen as more related to inflammatory alterations. Haapakoski and their colleagues found significant differences in levels of IL-1β for depressive patients when they limited the sample size with individuals aged more than 40 (Haapakoski et al., 2015). The mean age of patients with MDD in this study is 43.6 years, this may have restrained potential associations. Second, nearly 80% of groups in this study comprise women and several studies suggest that the relationship between depression and inflammatory molecules is stronger in men (Crosnoe, 2002; Elovainio et al., 2009; Vogelzangs et al., 2012). Therefore, gender differences could be another factor that causes the lack of significant associations. Third, 50% of our MDD sample were having their first episode of depression. It may be speculated that recurrent depressive episodes cause more inflammatory burden throughout a lifetime. Fourth, the lack of evidence for IL-1β could also partly be explained with methodological issues, conventional ELISA approaches might have difficulties detecting very low concentrations of interleukins. Lastly, the different patterns of the associations for IL-1β, NLRP3, ASC and NEK7 in this study could be justified by taking various biological pathways involving into account. The production of IL-1β could be stimulated by different inflammasomes like caspase-1 as mentioned above (Mathur et al., 2017). The recent study which investigates the anti-inflammatory effects of clomipramine showed that IL-1β expression was decreased by clomipramine. However, the drug did not have any impact on the NLRP3 expression (Gong et al., 2019).

There is only one clinical study that examined the correlations of NLRP3 inflammasome and depression severity. No correlations for NLRP3 and caspase-1 were shown, but they established a positive correlation for IL-1β (Alcocer-Gómez et al., 2014). Our results did not demonstrate any correlation of IL-1β and depressive symptom severity, as it was revealed in another large cohort study (Vogelzangs et al., 2012). With the existing limited evidence, it seems there is no association between levels of inflammatory markers and depression severity. In terms of different depression subtypes, atypical depression was proposed to be more related to inflammatory changes rather than melancholic depression (Lamers et al., 2013; Rothermundt et al., 2001), although it was not supported with later findings (Veltman et al., 2018). Due to the low number of patients with atypical MDD (n=6) in this study, no analysis was performed based on the depression subtype. We did not find any correlations of inflammatory molecules and the atypia index; yet, this index should not be recognized as a full-blown atypical depression.

The present study has several strengths. Participants were evaluated by experienced clinicians and diagnoses were made up based on objective structured interviews. Also, we excluded people on psychotropic medication which is advantageous as these medications affect inflammatory parameters including NLRP3 inflammasome (Alcocer-Gómez et al., 2017; Du et al., 2016). Furthermore, relatively bigger sample size was used compared to other few studies in the literature and NEK7 was evaluated for the first time within the context of MDD in a clinical study. On the other hand, the present study has some limitations. The cross-sectional design of the study does not allow to make any causal inferences for NLRP3 inflammasome and MDD. Inflammatory mechanisms are known to be affected by numerous biological and psychological factors and many of them could not be adjusted. It should always be acknowledged that presented associations may be partly driven by countless other determinants. The sample size and female dominance of the samples limit the generalizability of the findings.

In conclusion, the findings from this study align with the existing evidence that supports the role of the NLRP3 inflammasome complex in the pathogenesis of MDD. Yet, these results need to be replicated in larger sample sizes in light of future pre-clinical research focused on the biology of NLRP3 inflammasome.

## Data Availability

All data produced in the present study are available upon reasonable request to the authors.

## Authorship contribution statement

FÖ, BTO, YO, TA, EOA, and ND designed the study. FÖ and BTO collected the data; FÖ, TY, and BE analyzed the data. FÖ drafted the first manuscript; all the authors revised the manuscript for important intellectual content. All the authors approved the final version of the article.

## Notes

### Competing Interest Statement

The authors have declared no competing interest.

### Funding Statement

This study was funded by a grant from the Scientific and Technological
Research Council of Turkey (TUBITAK) (project number: 218S271).

### Author Declarations

Ethics committee/IRB of Dokuz Eylul University School of Medicine gave ethical approval for this work.

